# Signal-to-noise evaluation of dynamic versus static ^18^FDG-PET in focal epilepsy via Bayesian regional estimated signal quality analysis

**DOI:** 10.64898/2026.04.12.26350712

**Authors:** Mark Quigg, Pavel Chernyavskiy, William Terrell, Racheal Smetana, Thomas Evulathingal Muttikal, Megan Wardius, Bijoy Kundu

## Abstract

**Background and Purpose:** 2-[^18^F] fluoro-2-deoxy-D-glucose positron emission tomography (static PET) has mixed specificity and sensitivity in targeting epileptic zones in the noninvasive stage of epilepsy surgery evaluations. We compared the signal quality of static PET compared to a method of interictal dynamic PET (iD-PET).

**Materials and Methods:** We calculated the signal quality of static PET and iD-PET obtained from a cohort of patients with focal epilepsy. We developed a Bayesian regional estimated signal quality (BRESQ) technique to objectively compare signal-to-noise ratios (SNRs) by region of interest (ROI) within subjects.

**Results:** Adjusted for ROI size and neighboring regions, iDPET was superior to sPET with probability >95% in 8/36 regions; >90% in 21/36 regions; >80% in 29/36 regions. The top five regions with the largest adjusted SNR differences (greatest magnitude of iDPET superiority) were the Temporal Mesial (Left and Right), Occipital Lateral (Left and Right), and the Left Frontal Inferior Base.

**Conclusions:** We found that iDPET yielded a superior SNR in most ROI. BRESQ offers a scalable and generalizable method to quantify signal quality between brain mapping modalities.

## 1 Introduction

The identification of brain regions with impaired glucose metabolism with the use of ^18^F-fluorodeoxyglucose positron emission tomography (^18^FDG-PET) is a widely accepted method of identification of an hypometabolic epileptic zone in the evaluation for possible epilepsy surgery in patients with drug resistant epilepsy. However, standard static ^18^FDG-PET (sPET) has limitations. A recent, large, multicenter retrospective study found that sPET was normal in 48% of cases ^1^ of surgical candidates.

One reason why sPET may not identify a hypometabolic epileptic zone is that the ultimate destination of ^18^FDG, as a glucose mimic, is to collect where the greatest density of oxidative metabolism occurs, the mitochondria. These cluster at points of high energy use, the presynaptic region. Since uptake results from both synaptic density and synaptic activity, epileptic zones without clear structural lesions may not show up as hypometabolic because mitochondria density may not differ from tissue outside of the epileptic zone 2-4. Therefore, measures of metabolic rate may improve the identification of a metabolically-impaired epileptic lesion.

Our group developed a toolkit of acquisition and processing of dynamic ^18^FDG-PET that evaluates the kinetics of glucose uptake **(Figure 1A)** rather than the absolute standardized uptake value ^5-9^. Since epileptic zones consist of regions with decreased metabolism (due to down-regulated neuronal activity, altered connectivity, or focal mitochondrial dysfunction 10-12), the kinetics of ^18^FDG uptake **(Figure 1B)** may be more sensitive than static measurements of uptake density. Our preliminary work demonstrated that dynamic PET (interictal dynamic PET, iDPET) consistently shows focal lesions in animal models ^7^ and in those patients focal epilepsy whose sPET is normal ^5^ **(Figure 1C)**.

**Figure 1.**
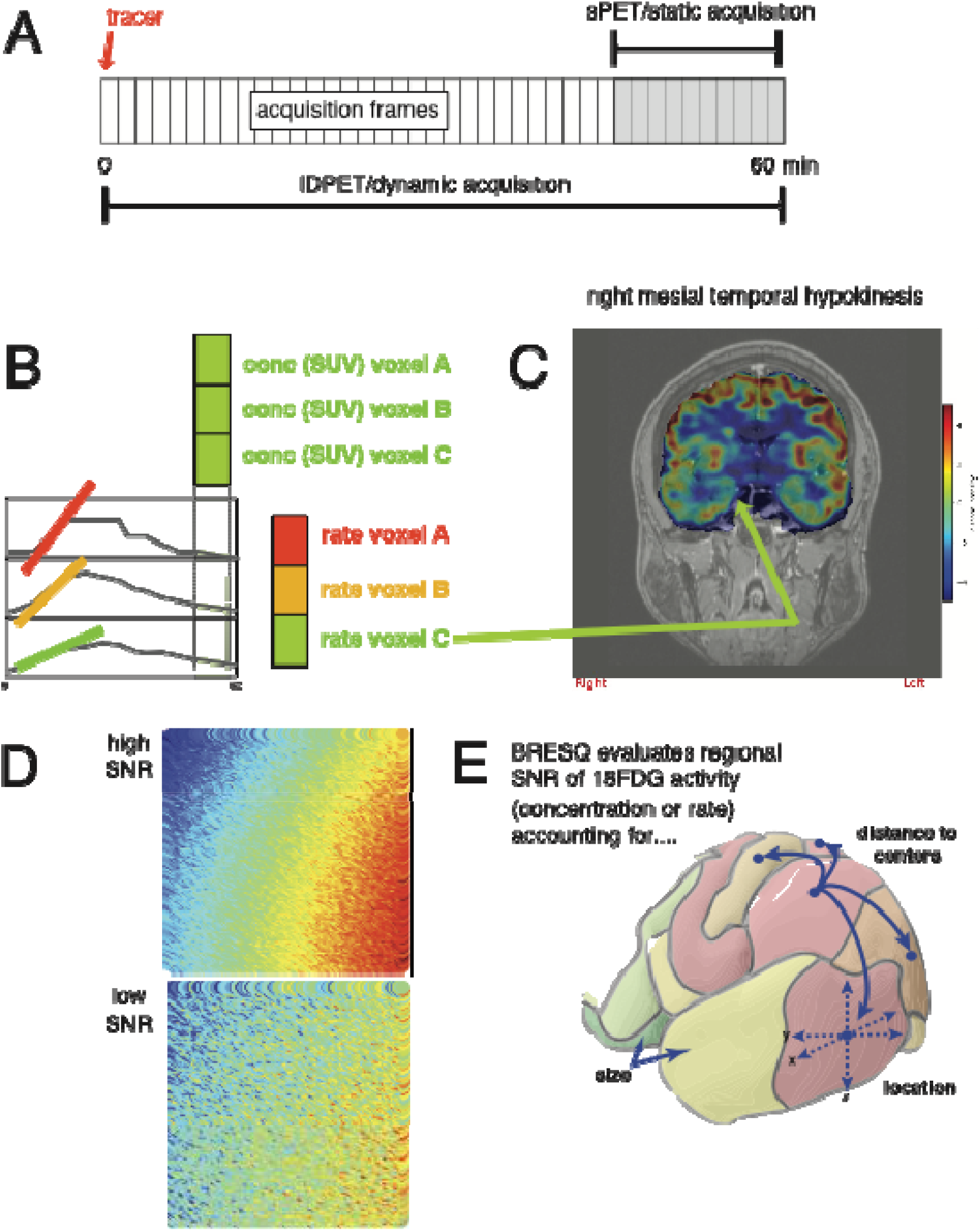
(A) Dynamic (iDPET) acquisition differs from static (sPET) ^18^FDG-PET in that (A) ^18^FDG is injected with the patient in the scanner, and acquisition recurs in separate frames for a one-hour total scan. Static PET images are extracted over a short series of frames. When traditional sPET is acquired, tracer is administer pre-scan, and after a waiting period to allow maximum uptake, a similar 10-15 minute acquisition is performed corresponding to the extracted sPET during a dynamic scan. (B) sPET standardized uptake values (SUV) sum up radiation concentration per voxel, whereas iDPET calculates the rate of ^18^FDG uptake per voxel. ^18^FDG uptake rates (known as k_i_, with an low uptake example marked by the green arrow), is then mapped on (C) a co-registered MRI in terms of z-scores calculated from the overall average ki, in this case demonstrating right mesial hypokinetics in a patient with drug-resistant mesial temporal lobe epilepsy. (D) A visual model demonstrating how an acquisition with high versus low signal-to-noise ratio may appear. The BRESQ protocol quantifies degree of signal-to-noise within and between images. (E) ki and concentration maps are processed via Bayesian Regional Estimated Signal Quality (BRESQ) technique to estimate regional signal-to-noise differences that account for patient differences, region location, interregion distance, and region size.

The critical initial step in the validation of iDPET is to determine differences between iDPET and sPET characteristics. One objective method is to evaluate the signal-to-noise ratio (SNR) ^13^ of the two imaging procedures. SNR is a unitless measure used to assess signal quality; higher SNR is interpreted as a stronger signal relative to the measured noise **(Figure 1D)**. The SNR is a widely accepted metric of comparative performance that has been used for brain MRI, PET, and EEG modalities ^14-16^.

In the present study, we compared the SNR of iDPET versus sPET in an epilepsy cohort to determine the better methodology of evaluating metabolic maps of resting state brain metabolism. The method with better SNR may then, in a subsequent validation study, more accurately discover epileptic lesions and surgical outcomes. We tested the hypothesis that kinetics of glucose uptake can better represent focal hypometabolism than static, absolute glucose uptake by virtue of higher SNR. We accomplished this comparison with an innovative Bayesian regional estimated signal quality (BRESQ) regression analysis of SNR that accounts for patient differences, regional brain differences, region location, and intra-brain correlations **(Figure 1E)**.

## 2 Methods

### 2.1 Subjects

In this IRB-approved, single center, observational study, consented patients were enrolled if they were 1) age > 17 years; 2) had at least 1 clinical-electrographic seizure on video-EEG; 3) were reviewed in the our Epilepsy Surgery Conference and determined to have focal DRE. iDPET was not used in diagnosis. Exclusions were 1) inability to obtain dFDG-PET without sedation; 2) diabetes mellitus; 3) psychogenic non-epileptic seizures or idiopathic generalized epilepsy; 4) weight >226kg; and 4) implantation of a neuromodulatory device.

### 2.2 Image acquisition and processing

The process of iDPET is detailed in our previous work^5, 8^ . In outline, dynamic acquisition occurred after intravenous injection of tracer before a 60-minute PET scan obtained in list-mode format **(Figure 1A)**. A high resolution T1-weighted MPRAGE MRI was obtained for co-registration. Subsequent processing was performed with custom tools that provided motion correction, image co-registration, transformation with a Montreal Neurological Institute ^17^ atlas, segmentation into 36 regions of interest (18 regions/side), calibration (model-corrected blood input function), and calculation of whole brain parametric kinetic uptake maps **(Figure 1B)**.

Data analysis was performed with z-score parametric mapping and segmented into regions of interest. Kinetic uptake maps for each patient were converted to voxel level z-score maps by normalizing to the whole brain mean and standard deviation within each patient (Figure 1C). sPET images were extracted from a 15 minute sample starting 45 minutes from study start; SUV maps were generated in a similar process to kinetic uptake maps. For SNR calculations, the brain was segmented into 36 regions of interest.

### 2.3 Bayesian regional estimated signal quality (BRESQ)

BRESQ is a novel process of quantifying SNR differences using a Bayesian regression that accounts for patient differences, region location, size, and intra-brain correlations to compare signal quality between mapping modalities. ^18^

In general, we define SNR as:

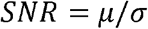

where *μ* = population mean and *σ* = population standard deviation. The sample SNR is estimated by the corresponding sample quantities: 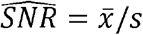, where the global standard deviation *s* serves as the measure of noise. Two SNR values can be compared by taking the difference 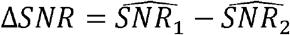.^22,26^

Here, we compared the signal quality of iDPET versus sPET by defining a new outcome variable *D* using the paired difference in SNR-based metrics for each brain region (*j* = 1, …,36) within patient (*i* = 1,…, N):

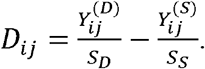

Above, 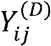 is the measured iDPET signal, 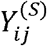 is the measured sPET signal, *S*_*D*_ is the standard deviation of iDPET signal across all patients and brain regions and *S*_*S*_ is the corresponding standard deviation of the sPET signal. Cases where *D*_*j*_ > 0 identifies brain regions where iDPET has superior signal quality to the sPET and vice-versa. In brain regions where *D*_*j*_ = 0, we will consider iDPET non-inferior to sPET (i.e., sPET is considered the standard of care).

Our regression is a hierarchical linear model that accounts for within-subject spatial correlation among brain regions and between-subject differences. To implement the spatial correlation, our team computed x, y, and z coordinates of each brain region centroid, unique to each patient. We performed our analysis under the Bayesian paradigm using standard code in the brms R package ^19^.

Preliminary exploratory analyses suggested: i) T distribution for the outcome due to kurtosis (heavy tails relative to a Normal distribution); ii) 3-dimensional anisotropic correlation that allows for differential spatial dependence in x, y, and z direction; and iii) the inclusion of brain region volume as a confounding variable, measured by the natural logarithm of the number of voxels (*logVox*). Thus, the statistical model was specified as follows:

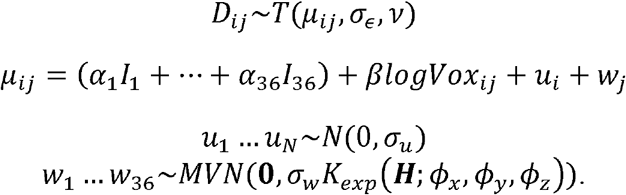

In the equation above, indicators *I*_1_ through *I*_36_ identify each of the 36 brain regions, *α*_1_ through *α*_36_ = corresponding fixed effects measuring mean iDPET vs. sPET differences, *u*_*i*_ = between-patient random intercepts, and *w*_*j*_ = within-patient brain region effects. Patient random intercepts (*u*_1_ … *u*_*N*_) were modelled as independent with *σ*_*u*_ as the subject random standard deviation. These effects reflect the joint contribution of all latent patient-specific factors (e.g., demographics, brain geometry, genetics) that may contribute to the outcome, aside from brain region and the size of each brain region. Finally, the random error standard deviation is measured by *σ*_*ϵ*_ and the *T* distribution degrees of freedom is measured by *v*.

To account for spatial correlation between brain regions (within patient) we used a stationary exponential correlation function 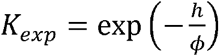, where *h* is the Euclidean distance between a pair of brain region centroids and *ϕ* controls how quickly the correlation decays over distance. For a given patient, the distance between any pair of brain regions *j* and *j*′ was computed as (*H*)_*jj*_′ = ‖ *h*_*j*_ − *h*_*j*_′ ‖, where *h* is the 3-dimension coordinate vector. The spatial standard deviation was measured by *σ*_*w*_ and spatial dependence in each of x, y, and z axis was measured by *ϕ*_*x*_, *ϕ*_*y*_, and *ϕ*_*z*_. The spatial effects (*w*_1_ … *w*_36_) were assumed to follow a mean-zero multivariate Normal distribution with covariance matrix *σ*_*w*_ *K*_*exp*_ (***H***; *ϕ*_*x*_, *ϕ*_*y*_, *ϕ*_*z*_).

Prior distributions – a statistical representation of prior knowledge, introduced prior to considering the data – were specified for all model parameters. These distributions were chosen to be weakly-informative for all parameters except for the spatial range. For all fixed effects (*α*_1_,…, *α*_36_, *β*) we used a Normal(0, 2.0), which specifies *apriori* a null effect on average, with mean SNR differences most likely to lie between -6 and 6. For the standard deviations (*σ*_*u*_, *σ*_*w*_, *σ* _*ϵ*_) we used a Half-Normal(0, 0.8) for the patient random intercepts, a Half-Normal(0, 0.5) for the spatial effects, and a Half-Normal(0, 0.8) for the error standard deviation. For the spatial range parameters (*ϕ*_*x*_, *ϕ*_*y*_, *ϕ*_*z*_) informative priors were chosen, as it is commonly recognized there is relatively little information to estimate these from the data: Inverse-Gamma(1.494, 5.823) for *ϕ*_*x*_, Inverse-Gamma(1.494, 7.056) for *ϕ*_*y*_, and Inverse-Gamma(1.494, 4.338) for *ϕ*_*z*_. For the degrees of freedom parameter *v* we used Inverse-Gamma(2, 0.1), which has infinite prior variance and is thus uninformative.

Because the analysis was Bayesian, we do not report p-values or Confidence Intervals. Instead, we report three Bayesian indices of effect existence: the 95% equal-tail Credible Interval (CrI), the posterior Probability of Direction (PD), and the Percent in Region of Practical Equivalence (% in ROPE) ^20^. The 95% CrI reflects the interval that contains the population parameter with 95% probability given the data. Formally, PD = *P*(*α*_*j*_ > 0) when the posterior median > 0 and the PD = *P*(*α*_*j*_ < 0) when the posterior median < 0. For our brain region effects (*α*_1_,…, *α*_36_), PD measures the probability of iDPET superiority for positive posterior medians and sPET superiority for negative posterior medians; PD is thus a key metric for quantifying SNR differences. Percent in ROPE is the probability that the posterior distribution reflects an arbitrarily-small and clinically negligible SNR difference, here assumed to be (-0.10, 0.10). Taken together, the strongest evidence of iDPET superiority is reflected by i) 95% CrI that excludes 0; ii) PD near 100%; and iii) % in ROPE near 0%. We note that because there are no p-values, we do not adjust post-hoc for multiple comparisons (e.g., via Bonferroni procedure) like one must do in a typical frequentist analysis.

We used 400 iterations as a warm-up and 5000 iterations to sample from the posterior, per each of the three initially overdispersed chains. Convergence was established through a visual inspection of MCMC chains and verifying that the Rhat statistics were near 1 for all parameters. The final model had satisfactory graphical posterior predictive checks showing no signs of over/underdispersion or bias^21^.

## 3 Results

### 3.1 Patients

Of the 32 patients enrolled and scanned, 30 were appropriate for analyses (**Table 1**). Two patients were excluded because of tracer quality or absent MRI data for co-registration.

**Table 1.** Patients (N=30) with drug-resistant focal epilepsy with dynamic and static ^18^FDG-PET.

|  |  | N | % |
| --- | --- | --- | --- |
| Sex | M | 14 | 47% |
|  | F | 16 | 53% |
| Race | White | 20 | 67% |
|  | Nonwhite | 10 | 33% |
| Ethnicity | NonHispanic | 30 | 100% |
|  | Hispanic | 0 | 0% |
|  |  | Mean | Standard deviation |
| Age (years) |  | 41.5 | 13.0 |
| Duration of epilepsy (years) |  | 14.8 | 13.5 |
| Seizure frequency (seizures/month) |  | 7.3 | 13.0 |

### 3.2 SNR iDPET versus sPET

Adjusted for patient differences, number of voxels, and correlation between brain regions, iDPET was superior to sPET with probability >95% in 8/36 regions; >90% in 21/36 regions; >80% in 29/36 regions **(Table 2)**. There were 5 regions where sPET was superior, with probabilities ranging from 76% (Right Frontal Opercular Broca) to 51% (Left Primary Sensory). The top five regions with the largest adjusted SNR differences (greatest magnitude of iDPET superiority) were the Temporal Mesial (Left and Right), Occipital Lateral (Left and Right), and the Left Frontal Inferior Base) (**Figure 2)**. On average, larger brain regions were associated with greater iDPET superiority in the SNR (*β*= 0.25 per 1 SD of voxels [95% CrI 0.02-0.49]) .

**Table 2.**
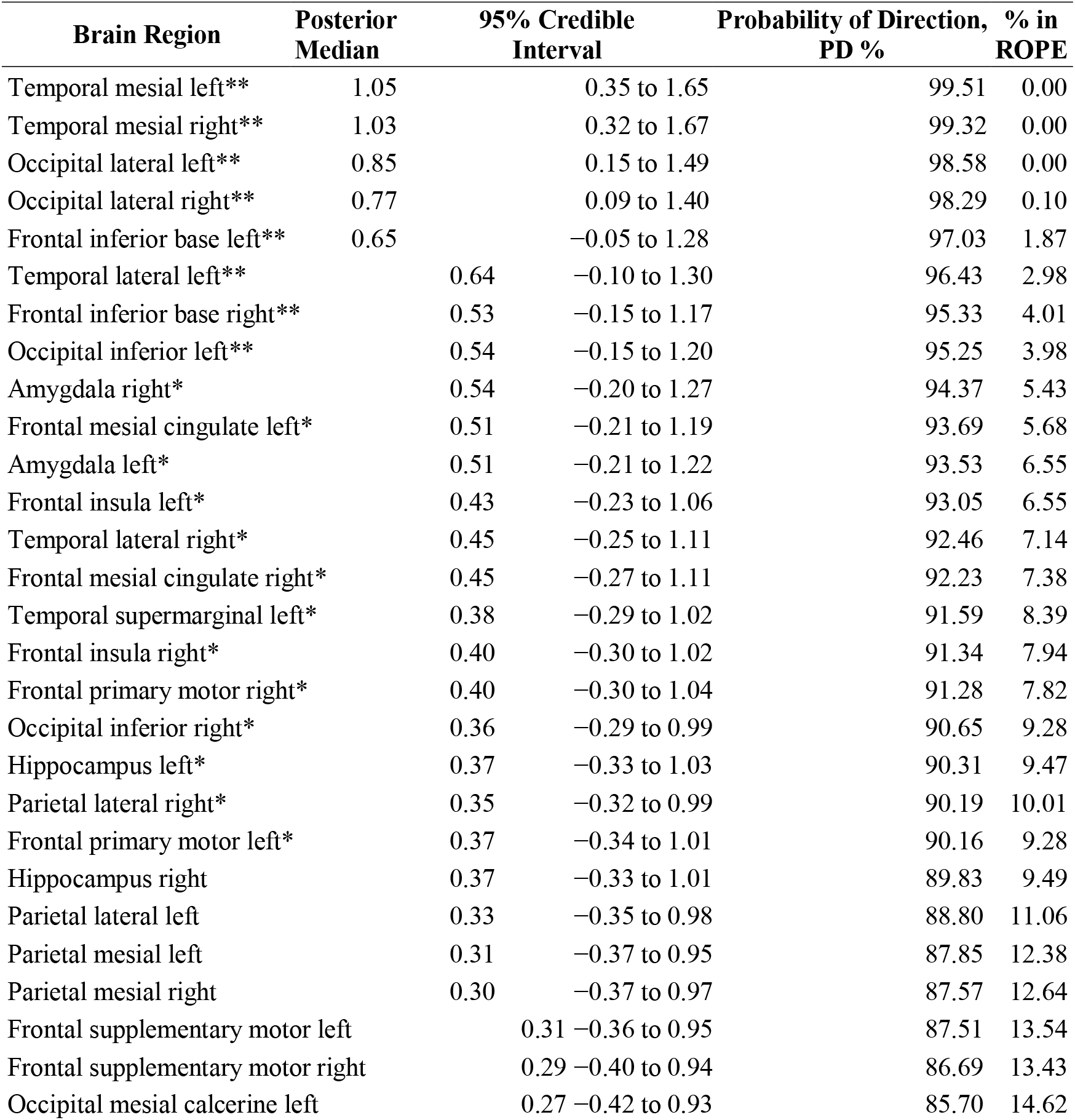
Model-adjusted iDPET vs. sPET differences in the SNR-based outcome by brain region ordered by the Probability of Direction (PD) (N = 30). ** indicates > 95% probability of iDPET superiority; * indicates > 90% probability of iDPET superiority. All estimates adjusted for patient differences, brain region volume, and correlation among brain regions. Region of Practical Equivalence (ROPE) is taken as [-0.10, 0.10].

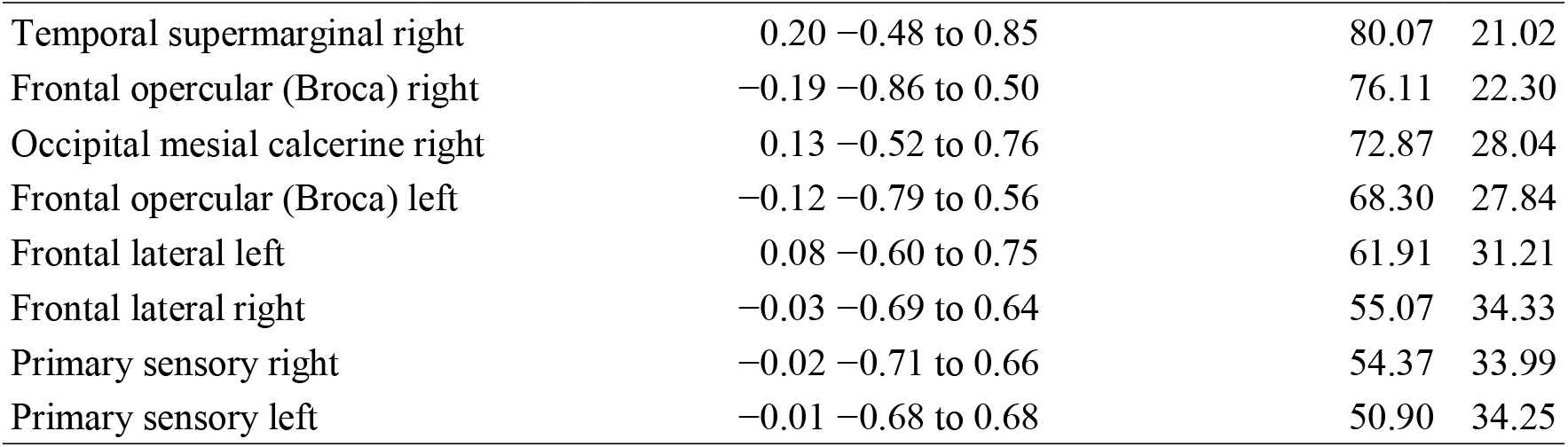

**Figure 2.**
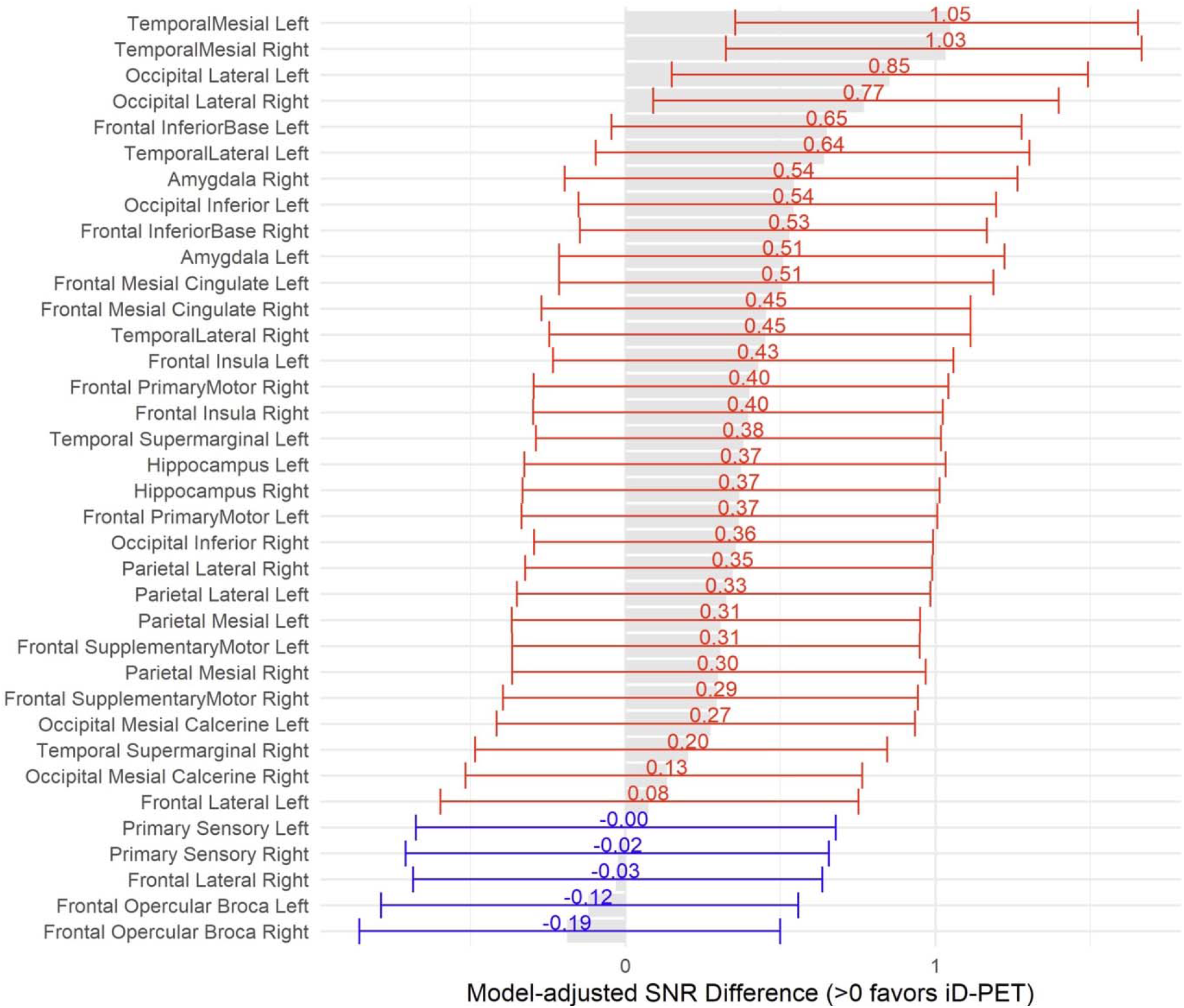
Bayesian regional estimated signal quality (BRESQ) comparisons between dynamic (iDPET) and standard (sPET) techniques in acquisition sof ^18^FDG-PET in 30 patients with focal drug resistant epilepsy. Brain regions of interested are ordered by adjusted SNR difference estimates. Error bars = 95% Credible Intervals; horizontal bars = Posterior Medians.

### 3.3 Between- and Within-Patient Effects

Between-patient effects – outside of brain region differences – accounted for 67% [95% CrI 33%-83%] of total variability in the outcome. Within-patient (between brain region) effects accounted for 10% [0%-55%] of total variability in the outcome. Correlation between brain regions was similar in the x and y directions (*ϕ*_*x*_ = 10.90 [1.26 − 53.89]; *ϕ*_*y*_ = 13.80 [1.54 − 67.16]) and weaker in the z direction (*ϕ*_*z*_ = 9.29 [0.92 − 44.09]) **(Figure 3)**.

**Figure 3.**
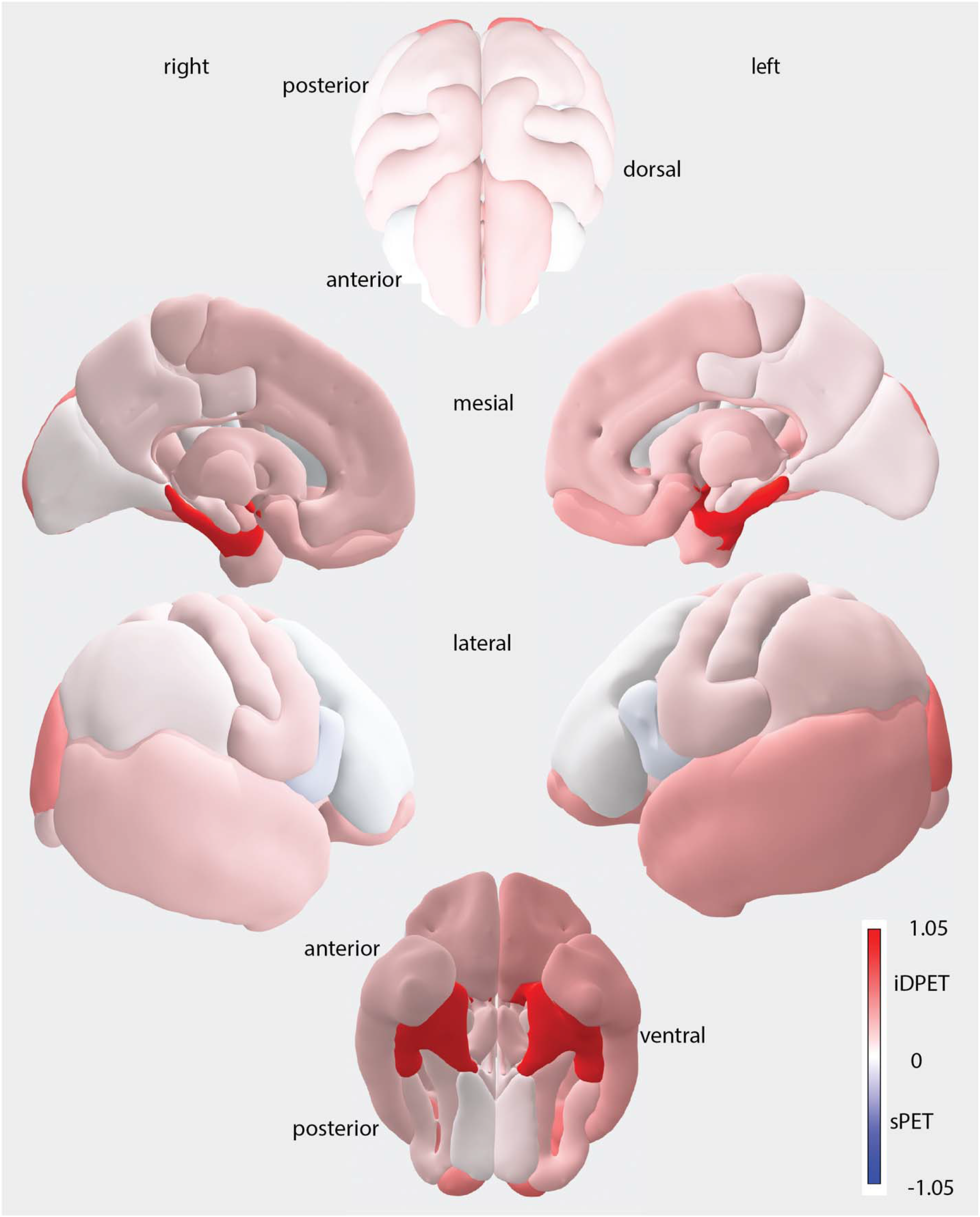
3D projections of the comparison of the BRESQ SNR differences of dynamic PET versus. traditional static PET divided into 36 surgically-relevant brain regions. In general, most regions showed a better signal quality of dynamic PET vs static PET.

## 4 Discussion

This analysis of the SNR characteristics of dynamic compared to static ^18^FDG-PET demonstrates that there is a regional distribution as well as an overall superiority of signal quality favoring iDPET. In general, iDPET shows better performance in the resolution of metabolic information in the limbic system, polar, and ventral regions of the cortex. Although a minority of regions have no clear difference between the SNR estimates, in no region was standard PET substantially superior to dynamic PET. Each comparison between iDPET and sPET was within each subject, so the location of the presumed epileptic zone did not affect comparisons. Dynamic PET may offer improved sensitivity in the identification of regions of focal hypometabolism by virtue of improved SNR.

Both biological and engineering factors probably play roles in improved signal quality. Dynamic PET captures both the rate of metabolism and the regional concentration of metabolism. Since regional glucose uptake varies with synaptic activity and synaptic density, capturing rate offers a second path of detection when density changes are subtle. An engineering advantage of kinetic models is that the calculation process takes into account the “blood partial volume”, the contribution of blood that contains ^18^FDG that flows through tissues and contaminates counts of regional uptake. In the case of dynamic acquisition, regional radiation accounts are adjusted for this background blood radiation but not accounted for in static scans ^24, 25^. The present study builds on earlier work in rodent brains ^22^ and animal models of movement disorders ^7^ and oxidative stress ^23^, human ambulatory metabolic studies ^24^, and brain cancer ^25^. Our first report in subjects with DRE detailed the findings of a cohort whose standard clinical sPET did not discover regions of hypometabolism ^5, 9^. iDPET, in contrast to sPET, identified regions of focal hypokinetic ^18^FDG uptake in all subjects. In short, we anticipate that patients with DRE will better demonstrate a focal region of hypometabolism uncontaminated by background blood flow regardless of the presence or absence of a gross structural lesion.

One area of innovation in this report is our use of a Bayesian spatial model (BRESQ) to quantify the differences in signal quality^18^. BRESQ was created to obtain more conservative statistical inferences relative to comparable frequentist models ^26^, gather more nuanced information about iDPET effects via Bayesian indices of effect existence ^20^, and to avoid making untenable asymptotic assumptions with small sample sizes. Because we used a robust statistical model that explicitly controls for brain region location, our approach is scalable and reproducible with a different set of brain segmentation techniques; in other words, BRESQ is a generalizable technique that can compare brain maps obtained from different modes. Accordingly, in future work we may apply this model to a different or greater number of regions of interest, or compare SNRs among a different set of brain imaging modalities (for example: structural MRI, functional MRI, or magnetoencephalography). With more regions, the assumption of spatial stationarity may be relaxed to allow the intra-patient spatial correlation to vary by brain region and potentially aid in EZ localization.

Our approach used sPET derived from each iDPET acquisition. The use of images derived from the same acquisition and from the same patient provide a strong counter for any effect that focal epilepsy may have on imaging. Our methodology, therefore, provides results that are relevant for healthy controls or those with other intrinsic brain lesions. Likewise, our analysis is reproducible under a different set of *a prori* assumptions by using a different set of prior distributions (e.g., uninformative or informative priors). If our analytic framework is used to compare imaging modalities with a greater knowledge base, informative prior distributions may be appropriate. ^27^

An unexpected finding was that brain regions differed in the degree to which iDPET offered improved signal quality over sPET. Although the correlation is not lock-step, we note that the regions with higher SNR in iDPET follow a ventral-to-dorsal gradient, with the higher SNR in the limbic system and other ventral regions. This gradient appears similar to the overall gradient in static metabolic labeling shown in a recent database of static ^18^FDG-PET brain images from 39 healthy controls ^28^. This report found that the temporal regions, both mesial and neocortical, demonstrated the least SUV activity. We surmise that the regions with higher SNR advantages are those in which background blood flow may contribute a greater proportion of regional radiation counts. The clinical implication is that those with EZs in regions with higher SNR improvements, such as temporal lobe epilepsy, may have the most to gain in use of iDPET in presurgical assessment. We anticipate that future work that will correlate iDPET findings with clinical findings aid in lateralization and localization of mesial temporal lobe epilepsy.

iDPET offers an advantage of scalability; it is potentially transportable to any facility with an appropriate PET scanner, a possible transformative method to allow current hardware to improve non-invasive localization. Our subsequent work has centered on means to automatize our process to a “turn-key” suite, including calibration steps (blood input function) performed with artificial intelligence methods ^8^. Our plan is to design trials that will calculate specificity and sensitivity of dFDG-PET to the “ground truth” of seizure freedom following epilepsy surgery.

A limitation was our division of regions of interest into 36 brain regions; future work will evaluate smaller, higher-resolution volumes. Some radiologists (and patients) may baulk at the one-hour scan times, but background work established this duration as optimal for rate calculations ^29, 30^. No patients failed the scan because of duration.

## Conclusions

In summary, this technical analysis established the regional distribution of signal quality of a functional neuroimaging technique in a cohort of patients with DRE. The technique of iDPET will be used in further trials to determine utility in epilepsy surgery. The BRESQ methodology may be used in future studies to rank localization modalities in reliability in relation to epilepsy surgery outcomes. Dynamic PET in general can be further explored where metabolic brain mapping can aid the characterization of other brain disorders.

## Data Availability

Data are available upon reasonable request to the authors.

## Abbreviations

